# Lesion sterilisation and tissue repair with chloramphenicol, tetracycline, zinc oxide-eugenol paste compared to zinc oxide-eugenol pulpectomy in primary molars: Two-year follow-up findings of a multicentre non-inferiority randomised clinical trial

**DOI:** 10.64898/2026.11.04.26367310

**Authors:** Pablo Silveira Santos, Tatiane Ramos, Gustavo da Costa Fernandes, Natália Matsuda de Oliveira, Aurélio de Oliveira Rocha, Filipe Colombo Vitali, Fabiola Fontes Galdino, Renata Pereira de Samuel Marques, Vera Lígia Vieira Mendes Soviero, Carla Miranda Santana, Tatiana Kelly da Silva Fidalgo, Mariane Cardoso, Fausto Medeiros Mendes

## Abstract

**Background:** Chloramphenicol, tetracycline, zinc oxide (CTZ) paste has been widely used, particularly in Latin America, in Lesion Sterilization and Tissue Repair (LSTR) procedures in primary molars. However, reports on its long-term efficacy are scarce.

**Aim:** This multicentre non-inferiority randomised trial compared the efficacy of LSTR using CTZ paste with conventional zinc oxide-eugenol (ZOE) pulpectomy in primary molars.

**Design:** 199 primary molars from children aged 3–9 years were randomised to the CTZ or ZOE groups. The primary outcome was clinical and radiographic success at 24 months. Clinical time, children’s behaviour, immediate discomfort and postoperative pain were also evaluated. The main analysis followed the intention-to-treat (ITT) principle, comparing success proportions with the Miettinen–Nurminen method, considering a prespecified non-inferiority margin of 15% (p < 0.05).

**Results:** Overall success rates in the ITT population were 64.3% for CTZ and 58.7% for ZOE, with a difference of 5.6% (95% CI: −9.3 to 20.4; *p*<0.01). Mean; standard deviation clinical time was significantly shorter in CTZ (36.3; 9.8 min) than in ZOE (63.0; 16.3 min) (*p*<0.01). Children’s behaviour, immediate discomfort and postoperative pain were similar between groups (*p*>0.05).

**Conclusion:** LSTR with CTZ paste was non-inferior to conventional ZOE pulpectomy after 24 months of follow-up.

## 1 Introduction

Maintaining primary teeth in a healthy, functional state until natural exfoliation is a fundamental objective of paediatric dental care, as premature tooth loss may compromise oral development, the integrity of the permanent successors, and children’s oral health-related quality of life.^1^ When the pulp becomes irreversibly inflamed or necrotic, pulpectomy is one of the possible management alternatives.^2,3^ Conventional pulpectomy involves chemomechanical preparation of root canals, followed by obturation with a resorbable and biocompatible filling material.^2,4,5^ Evidence from a randomised trial showed that pulpectomy leads to a better oral health-related quality of life compared to tooth extraction.^6,7^

Nevertheless, conventional pulpectomy in primary teeth presents several difficulties. Complex root canal anatomy, physiological or pathological root resorption, and the need to manage young children with variable levels of cooperation may all compromise treatment outcomes.^2,5^ To deal with these difficulties, simplified endodontic approaches have been proposed to reduce this complexity while preserving the primary tooth. Among them, Lesion Sterilisation and Tissue Repair (LSTR), which relies on disinfection of the root canals with an antibiotic paste without mechanical instrumentation, has emerged as a promising alternative.^8–10^

LSTR is based on the principle that adequate disinfection of the infected root canal system may be achieved without mechanical instrumentation. Instead, an antibiotic mixture is placed within the pulp chamber and at the entrances of the root canals to reduce the microbial load and promote tissue repair.^2,4,11^ The introduction of the LSTR concept and its biological rationale was established in the early 1990s.^8–10^ Since then, the technique has been investigated as a simplified alternative to conventional pulpectomy, particularly for managing necrotic primary teeth.

Several antibiotic formulations have been proposed for use in LSTR procedures. One of the earliest was CTZ paste, originally described in 1964.^12^ The formulation consists of chloramphenicol, tetracycline, zinc oxide, and eugenol as the vehicle. Following the development and dissemination of the LSTR concept, CTZ paste was adopted as one of the antibiotic formulations used in LSTR protocols.^12^ More recently, the triple-antibiotic paste known as 3Mix, comprising minocycline, metronidazole, and ciprofloxacin in a propylene glycol vehicle, has become one of the most extensively investigated antibiotic formulations in studies evaluating LSTR.^11^

Growing interest in LSTR was reflected in its inclusion as a treatment option in the 2020 guideline on non-vital pulp therapies in primary teeth published by the American Academy of Pediatric Dentistry (AAPD).^3^ According to the guideline, LSTR may be considered for teeth presenting preoperative internal or external root resorption or when short-term retention of a primary tooth, up to 12 months, is desirable as an alternative to extraction.^2,3^ Nevertheless, the certainty of the available evidence remains limited, and stronger recommendations depend on the availability of well-designed randomised clinical trials with representative samples and adequate follow-up periods.^2,3^

Overall, long-term evidence on the effectiveness of LSTR, whether using 3Mix or alternative antibiotic formulations, remains scarce.^13,14^ Furthermore, many previous studies investigating CTZ paste have been limited by small sample sizes and heterogeneous comparator groups, which restricts the interpretation and applicability of their findings.^15–17^ Only one randomised clinical trial has evaluated the long-term clinical and radiographic success of LSTR using CTZ paste over a 36-month follow-up period.^18^ However, the authors tested a two-sided hypothesis and found no statistically significant difference between groups, yielding inconclusive evidence regarding either superiority or non-inferiority of the technique compared with conventional root canal treatment involving instrumentation.

To address this gap, we conducted a multicentre randomised clinical trial to compare the efficacy of LSTR using CTZ paste with conventional pulpectomy involving manual root canal instrumentation and zinc oxide–eugenol (ZOE) filling in primary molars. We hypothesised that the success rate of LSTR would be non-inferior to that of conventional pulpectomy after 24 months of follow-up, considering a pre-specified non-inferiority margin.

## 2 Material and Methods

The present report was prepared in accordance with the guidelines from the Consolidated Standards of Reporting Trials (CONSORT 2025 Statement)^19^ and the CONSORT extension for Reporting of Noninferiority and Equivalence Randomised Trials.^20^

### 2.1 Patient and public involvement

Because the study was planned during the COVID-19 pandemic and involved paediatric participants, the study had no formal patient or public involvement in the design, conduct, or reporting of the trial.

### 2.2 Trial design and ethical considerations

This was a multicentre, non-inferiority, controlled trial, with balanced allocation (1:1) to two parallel groups. The study protocol was approved by the local ethical committees from the enrolled centres: School of Dentistry of the University of São Paulo (FOUSP), Federal University of Santa Catarina (UFSC), Rio de Janeiro State University (UERJ) and Iguaçu University (UNIG), under the approval number 42602820.0.1001.0121. In addition, the study protocol was prospectively registered in ClinicalTrials.gov (Identifier: NCT04942158) and published elsewhere.^21^ Children were enrolled in the study only after providing written assent or, if they could not read or write, verbal assent, and after their legal guardians signed an informed consent form.

### 2.3 Changes to trial protocol

The first version of the study protocol anticipated the participation of four research centres. However, because of difficulties with participant recruitment and enrolment, one centre (UNIG) was discontinued during the study, and the sample size calculation was revised to reflect this change. We recorded all protocol amendments in the public trial registry when they were made. In addition, the secondary outcomes related to participants’ Oral Health-Related Quality of Life and the economic analysis will be presented in separate publications.

### 2.4 Trial setting

This multicentre study was conducted in university-based paediatric dental clinics providing secondary care at three Brazilian dental schools in São Paulo (FOUSP), Florianópolis (UFSC), and Rio de Janeiro (UERJ). Participants were recruited between October 2021 and March 2023 and were subsequently followed for 24 months after treatment.

### 2.5 Eligibility criteria

Children aged 3 to 9 years presenting a primary molar exhibiting clinical or radiographic signs of pulp necrosis were eligible for inclusion. Children with special healthcare needs, immunocompromising conditions, known allergies to the study medications, or uncooperative behaviour during the initial clinical examination were excluded.

Tooth-related inclusion criteria comprised spontaneous pain, fistula, swelling, and a history of these signs or symptoms. Radiographically, primary molars with furcation radiolucency not involving the follicle of the permanent successor tooth, retaining at least two-thirds of their root length, and no pathological internal root resorption or perforation of the pulp chamber floor were considered eligible. Teeth with non-restorable crowns, uncontrolled bleeding following pulp chamber access, pulp canal obliteration, significant loss of bone support, or excessive mobility were excluded.

### 2.6 Intervention and comparator

Participants in both groups received the same local anaesthesia protocol. Initially, 20% benzocaine gel (Benzotop, Nova DFL, Rio de Janeiro, Brazil) was applied to dried mucosa for two minutes to provide topical anaesthesia. After administering local anaesthesia with lidocaine 2% containing epinephrine 1:100,000 (Alphacaine, Nova DFL, Rio de Janeiro, Brazil) and isolating the rubber dam, the pulp chamber was accessed using an appropriately sized round diamond bur (KG Sorensen, São Paulo, Brazil) in a high-speed handpiece with water cooling. The access cavity was then refined with a non-cutting tip Endo-Z bur (Dentsply Maillefer, Ballaigues, Switzerland). After locating the root canal orifices, the sealed allocation envelope was opened to determine the assigned treatment.

In the CTZ (experimental group), the pulp chamber was irrigated with 5 mL of 1% sodium hypochlorite (NaOCl). After drying the pulp chamber with sterile cotton pellets, CTZ paste was placed at the root canal entrances using a No. 5 dental explorer, followed by gentle pressure with sterile cotton pellets moistened with 0.9% sterile saline solution. The CTZ paste (Fórmula & Ação, São Paulo, Brazil) was prepared by mixing 62.5 mg of chloramphenicol, 62.5 mg of tetracycline, and 125 mg of zinc oxide with one drop of eugenol.

In the ZOE (control group), the working length (WL) was determined radiographically and established 1 mm short of the radiographic apex. Root canals were instrumented using 21-mm stainless steel endodontic manual K-files (Dentsply, Ballaigues, Switzerland), ranging from sizes #15 to #40. Irrigation with 2 mL of 1% NaOCl was performed after using each file. The first file to bind at the WL was designated as the initial apical file (IAF). Canal preparation was then completed using at least two successively larger files to the same working length, with a master apical file of at least size #35. Final irrigation was performed using 10 mL of 17% ethylenediaminetetraacetic acid (EDTA) for 3 minutes, followed by 0.9% sterile saline solution. The root canals were dried with absorbent paper points and filled with ZOE paste (Biodinâmica, Rio de Janeiro, Brazil). The filling material was inserted into the root canals using a Lentulo spiral operated in a low-speed handpiece to a depth of 2 mm short of the WL. The ZOE paste was prepared by mixing 250 mg of zinc oxide with five drops of eugenol.

Both groups received the same restorative protocol. A thin layer of glass ionomer cement (Encapsulated Riva Self Cure, SDI Ltd., Victoria, Australia) was applied over the filling material to prevent direct contact with the composite resin. Subsequently, the teeth were restored with a bulk-fill resin composite (Filtek Bulk-Fill Posterior Restorative, A1 shade; 3M ESPE, Minnesota, United States of America) using a three-step etch-and-rinse adhesive system (Adper Scotchbond Multipurpose, 3M ESPE, Minnesota, United States of America), according to the manufacturer’s instructions. All treatment procedures were completed in a single visit.

### 2.7 Outcomes

#### 2.7.1 Primary outcome

The primary outcome was the success of the endodontic treatment at the 24-month follow-up, assessed through clinical and radiographic evaluations. Clinical success was defined as the absence of pain, fistula, swelling, or pathological mobility, alongside the presence of periodontal health or physiological exfoliation. Radiographic success was defined by: (1) reduction or stabilization of pre-existing furcation radiolucency, (2) root resorption compatible with the expected stage of physiological exfoliation, (3) absence or stabilization of pathological root resorption, and (4) maintenance of an intact coronal restoration providing an adequate seal. The presence of any clinical or radiographic sign of failure was considered indicative of treatment failure.

#### 2.7.2 Secondary outcomes

The clinical operating time was recorded in minutes and seconds using a digital chronometer. An independent assistant recorded the total time required to complete each procedure, from the topical anaesthesia to the removal of the rubber dam isolation after tooth restoration.

The children’s behaviour during the procedures was evaluated using the Frankl Behaviour Rating Scale.^22^ This ordinal scale classifies behaviour into four categories: definitely negative (score 1), negative (score 2), positive (score 3) and definitely positive (score 4). Calibrated assessors (one from each centre) classified the children’s behaviour in the period after local anaesthesia until the removal of rubber dam isolation.

Following the interventions, children’s self-reported immediate discomfort was assessed using the Facial Image Scale.^23^ This ordinal scale comprises five facial expressions representing a spectrum from a very sad and crying face (score 5) to a very happy and smiley face (score 1). An independent assessor presented the scale to the children and inquired: “Which of these faces best represents how you feel after your tooth treatment?”.

At 48 hours post-intervention, postoperative pain, swelling, and analgesic consumption were assessed through a telephone interview with the child’s parent or caregiver. The assessment included the following questions: “Has your child reported pain in the treated tooth (yes/no)?”, “Have you noticed any swelling in the treated area (yes/no)?”, and “Did your child require any analgesic medication (yes/no)?”

### 2.8 Harms

At the 48 hours post-intervention telephone interview, parents and caregivers were also invited to report any additional discomfort or adverse events experienced by the children following treatment.

### 2.9 Sample size

The sample size was calculated using the Sealed Envelope® estimator (https://www.sealedenvelope.com/power/binary-noninferior/). An initial sample of 114 participants was calculated, assuming a success rate of 92%, a non-inferiority margin of 15%, a significance level of 5%, and 90% power. Adding 20% considering possible losses of follow-up and 60% due to the multicentre design (20% to each additional centre), the required sample size was 218 participants (109 in each group). This sample size was revised on two occasions due to slower-than-anticipated participant recruitment and the discontinuation of one study centre. Following these adjustments, the minimum required sample size was recalculated as 182 participants (91 per group).

The non-inferiority limit was determined based on approximately 50% of the difference in treatment success rates (27%) between LSTR technique and conventional pulpectomy in primary teeth reported in a previous systematic review.^2^

### 2.10 Sequence generation, allocation concealment and implementation

Children were randomly allocated to one of the two study groups using permuted randomisation with block sizes of 2, 4, and 8. Randomisation procedures were stratified by the study centres. An independent assistant generated the allocation sequence using a computer-generated randomisation list created on the Sealed Envelope® website. To ensure allocation concealment, an independent assistant prepared sequentially numbered, opaque, sealed envelopes for each study centre, and the envelope was opened only after pulp chamber access,

### 2.11 Blinding

This was an open-label trial. Considering the inherent differences between the interventions under investigation, blinding of operators, participants, and outcome assessors was not feasible.

### 2.12 Training and calibration exercises

Before patient recruitment, one operator from each study centre was trained in both endodontic techniques (LSTR and conventional pulpectomy technique). The training included theoretical classes on the clinical procedures of both techniques, followed by a pilot test of the clinical protocol in five children not included in the study.

One assistant from each centre was trained and calibrated to use the Frankl Behaviour Rating Scale to assess children’s behaviour during treatment. A paediatric dentist with a master’s degree in Dentistry and previous experience with the scale served as the benchmark examiner for calibration. Ten videos of dental appointments were evaluated, in which the assistants classified the children’s behaviour. This activity was conducted twice, with a 14-day interval between them, and inter-and intra-examiner agreement indices were calculated using the weighted *Kappa* coefficient. The intra-examiner Kappa values obtained ranged from 0.852 to 1.000, while the inter-examiner values ranged from 0.868 to 1.000.

Furthermore, three assistants were trained to conduct the clinical follow-up examinations, and one examiner, a specialist with a PhD in Endodontics, was trained and calibrated to perform radiographic evaluation. Radiographs of 10 cases treated with both endodontic techniques tested in this study were evaluated and compared with the assessment of a Paediatric Dentistry Professor, who served as the benchmark examiner. Similarly, the calibration process was conducted in two stages, with a 14-day interval between them, and inter-and intra-examiner agreement indices were calculated using the *Kappa* coefficient. The inter-examiner agreement index obtained was 0.783, while the intra-examiner agreement index was 1.000.

### 2.13 Statistical methods

Initially, descriptive analyses were conducted to present the number and frequency of participants randomly allocated to each treatment group, as well as the number of participants followed up to 24 months and the dropouts.

The primary outcome was analysed under a non-inferiority framework in the full intention-to-treat (ITT) population. We handled missing outcome data using multiple imputation by chained equations (MICE) with logistic regression, incorporating centre, treatment group, age, tooth type, and the presence of abscess or fistula, interradicular lesion, and external resorption as predictors. Non-inferiority was assessed using the Miettinen– Nurminen method stratified by centre. Estimates obtained from the 50 imputed datasets were pooled using Rubin’s rules. *P*-values and 95% confidence intervals (95%CI) for the risk difference were generated considering a prespecified non-inferiority margin of 15%.

Sensitivity analyses were also conducted. First, we performed a modified ITT analysis based on complete cases, excluding participants from the centre that was dropped (UNIG, *n* = 20). For this analysis, missing data from children not followed up to 24 months were imputed using the same MICE procedure described above, and non-inferiority was again evaluated using the Miettinen–Nurminen method.

We performed a second sensitivity analysis in the per-protocol population, restricted to subjects who completed the 24-month follow-up. In this analysis, the experimental group was considered non-inferior when the lower limit of the 95% CI for the risk difference did not cross the non-inferiority margin and the *p*-value from the Miettinen–Nurminen method stratified by centre was below 5%. Finally, a Cox regression analysis was conducted to assess non-inferiority among children with at least one follow-up assessment. In this analysis, hazard ratios (HRs) and their 95% CIs were estimated, and a non-inferiority margin of 1.57 for the HR was adopted, which corresponds to the 15% risk-difference margin defined for the primary endpoint.

Subgroup analyses were conducted in the full ITT population based on the following baseline characteristics: centre, sex, age (categorised as 3–5 years or >5 years), tooth type, and presence of an abscess or fistula. For each subgroup, risk differences and corresponding 95% CIs were derived using the Miettinen–Nurminen method within each of the 50 imputed datasets, and then combined using Rubin’s rules to obtain pooled estimates. We presented the final estimates in a forest plot.

Clinical and radiographic success (secondary outcomes) were separately analysed under a non-inferiority framework, using the same methodological approach as for the primary endpoint (overall success). Other secondary outcomes were compared between the groups in the full ITT population using two-tailed hypothesis tests. Dichotomous outcomes (occurrence of pain, oedema, or need for analgesics) were imputed using MICE (50 imputations), and groups were compared using a centre-stratified Wald test within each imputed dataset, with pooled estimates derived through Rubin’s rules.

Descriptive analyses were conducted using StataSE 15.0 (StataCorp LLC, College Station, USA), and other analyses were conducted in the R platform (R 4.5.1, The R Foundation for Statistical Computing, Vienna, Austria). The level of significance was set at 5% in all analyses. The anonymised dataset (https://repositorio.uspdigital.usp.br/handle/item/867), analysis codes (https://repositorio.uspdigital.usp.br/handle/item/888) and full statistical report (https://repositorio.uspdigital.usp.br/handle/item/887) are available in the public data repository of the University of São Paulo.

## 3 Results

A total of 402 children were assessed for eligibility, of whom 199 were included and randomised into two study groups: 100 to the CTZ and 99 to the ZOE. Two participants did not receive the allocated intervention because perforation at the entrance of a root canal was identified after randomisation. Participants were initially recruited from four study centres; however, one centre did not complete the study due to challenges with recruitment and participant recall, resulting in the loss of all enrolled participants from that centre (*n* = 20). Thus, 156 of the 199 randomised participants completed the 24-month follow-up (76 in the ZOE and 80 in the CTZ). The overall attrition rate was 21.6%, corresponding to 43 participants. Details concerning participant enrolment, allocation, and follow-up are presented in the study flow diagram (Figure 1).

**Figure 1.**
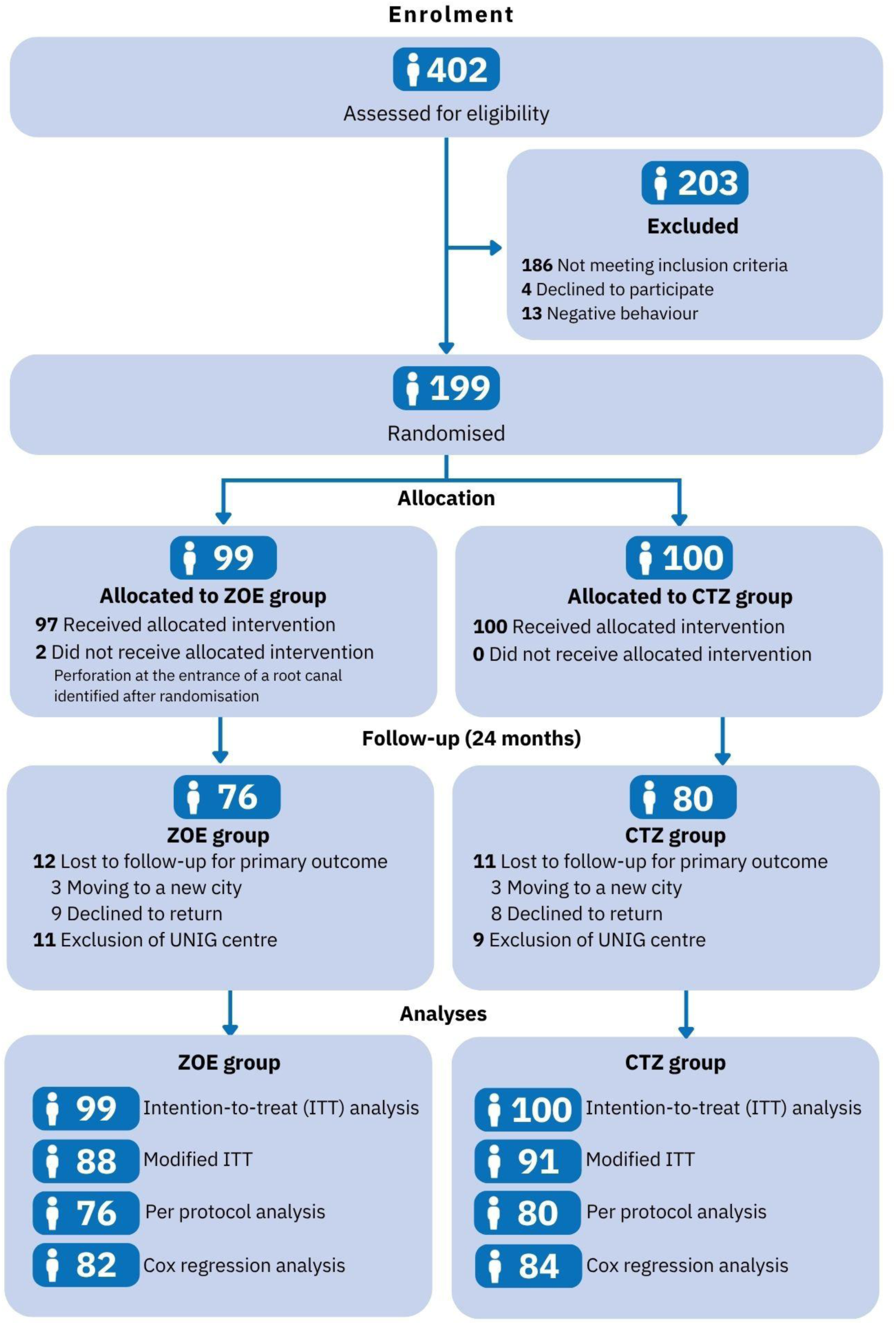
CONSORT flow diagram presenting an overview of patient enrolment, allocation and follow-up processes.

The baseline characteristics of the sample are summarised in Table 1. The mean (standard deviation - SD) age of the participants was 5.8 (1.4) years, with a predominance of boys (56.8%). Most included teeth were second primary molars (61.8%) in the mandibular arch (65.8%). Nearly half of the teeth had a fistula or abscess (48.7%), and most showed interradicular radiolucency (84.4%). Furthermore, most teeth did not present external root resorption at baseline (85.4%).

**Table 1.**
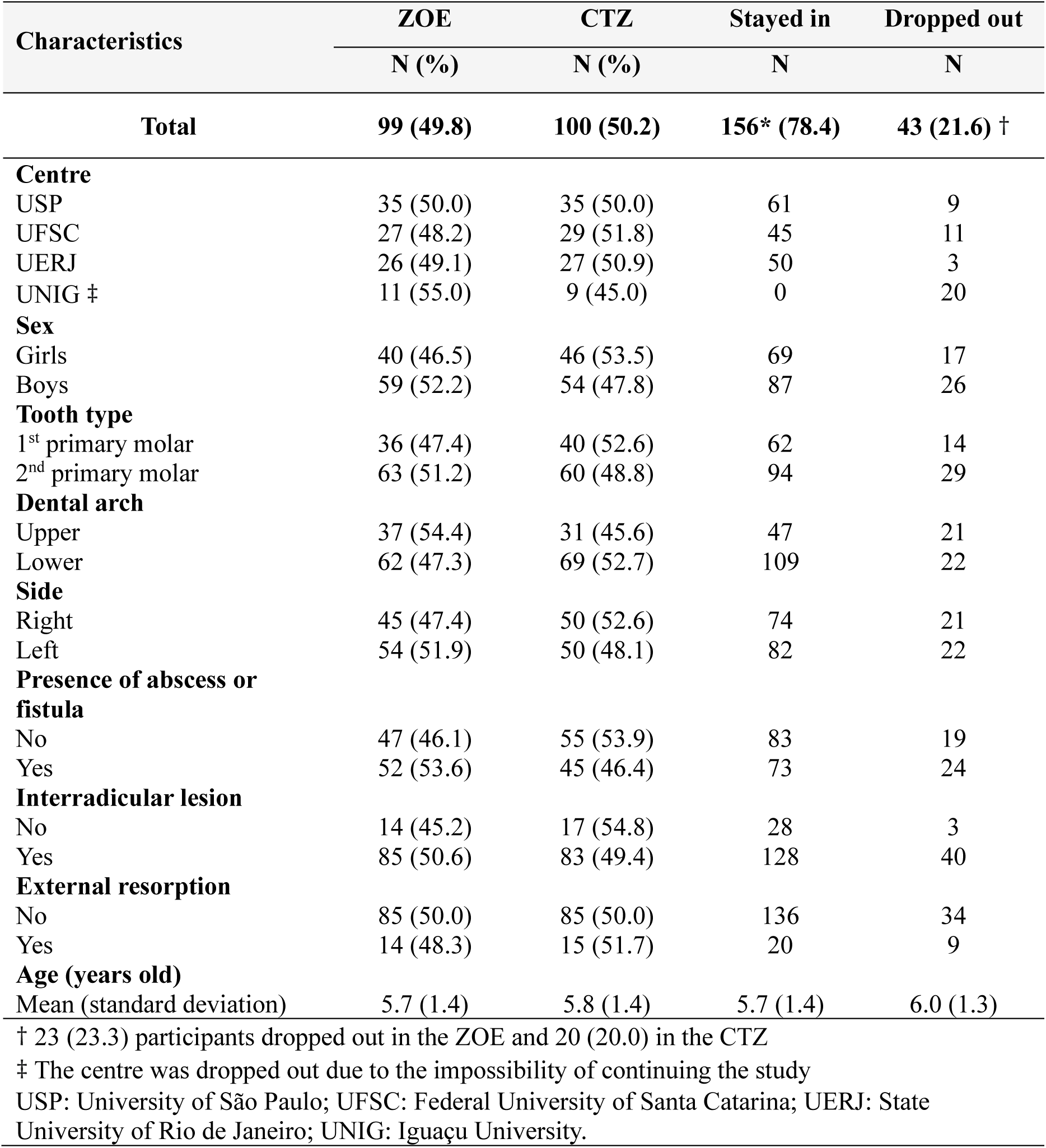
Baseline characteristics of participants and randomised teeth according to allocated group.

Table 2 shows the main analysis considering the primary endpoint. Among children who completed the 24-month follow-up (per-protocol population, *n =* 156), treatment success was observed in 45/76 teeth (59.2%) from ZOE, and in 51/80 teeth (63.8%) from CTZ (Table 2). In the ITT population (prespecified primary analysis), the non-inferiority of the CTZ was demonstrated, as the lower limit of the 95% CI for the difference in success rates (CTZ − ZOE) was-9.3%, remaining above the non-inferiority margin of-15.0% (Figure 2 and Table 2).

**Figure 2.**
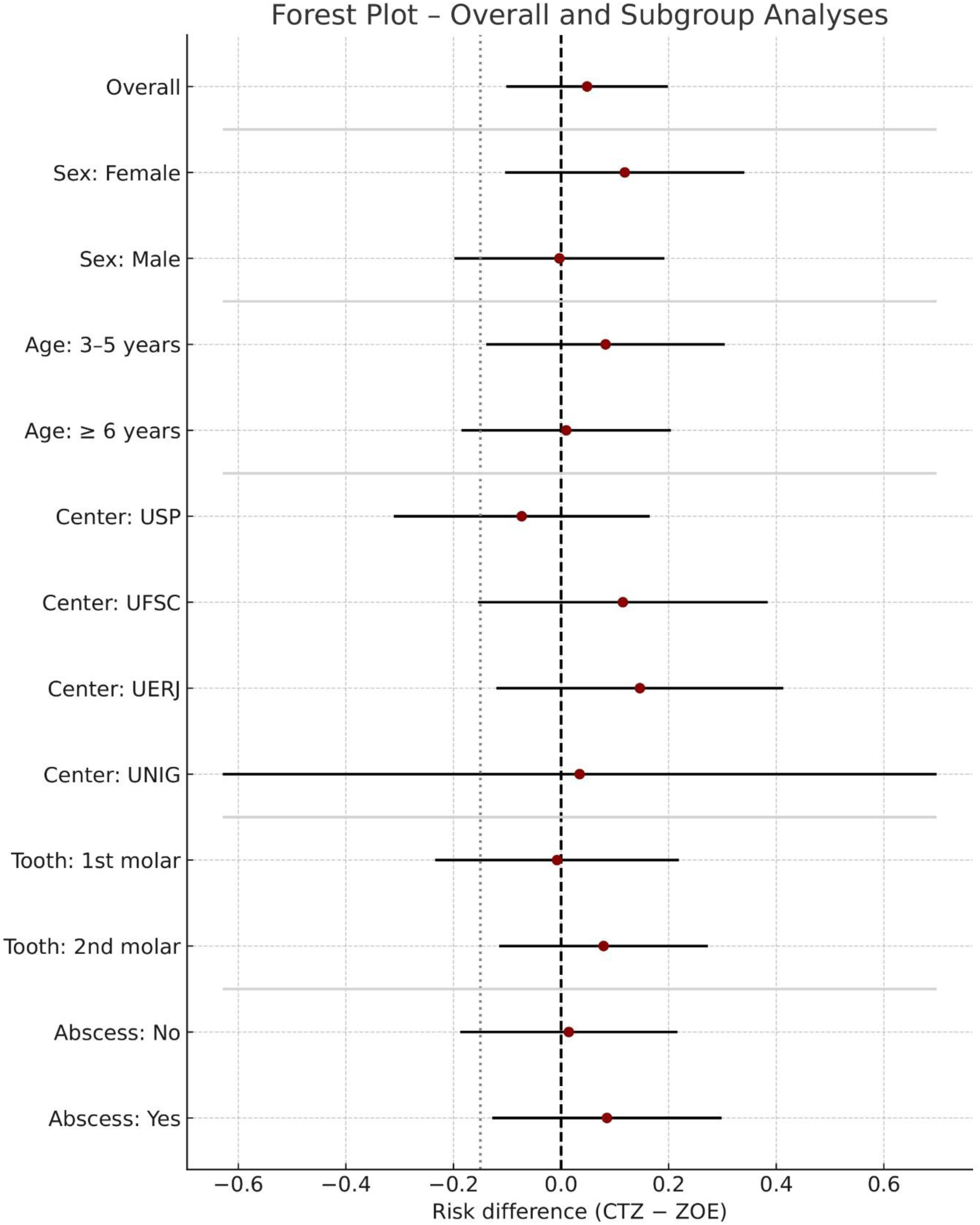
Differences in overall success in the intention-to-treat population and subgroup analyses. Dots represent the absolute differences, and horizontal lines represent 95% confidence intervals. The vertical dotted line indicates the prespecified non-inferiority margin. USP – University of São Paulo; UFSC – Federal University of Santa Catarina; UERJ – Rio de Janeiro State University; UNIG – Iguaçu University.

**Table 2.**
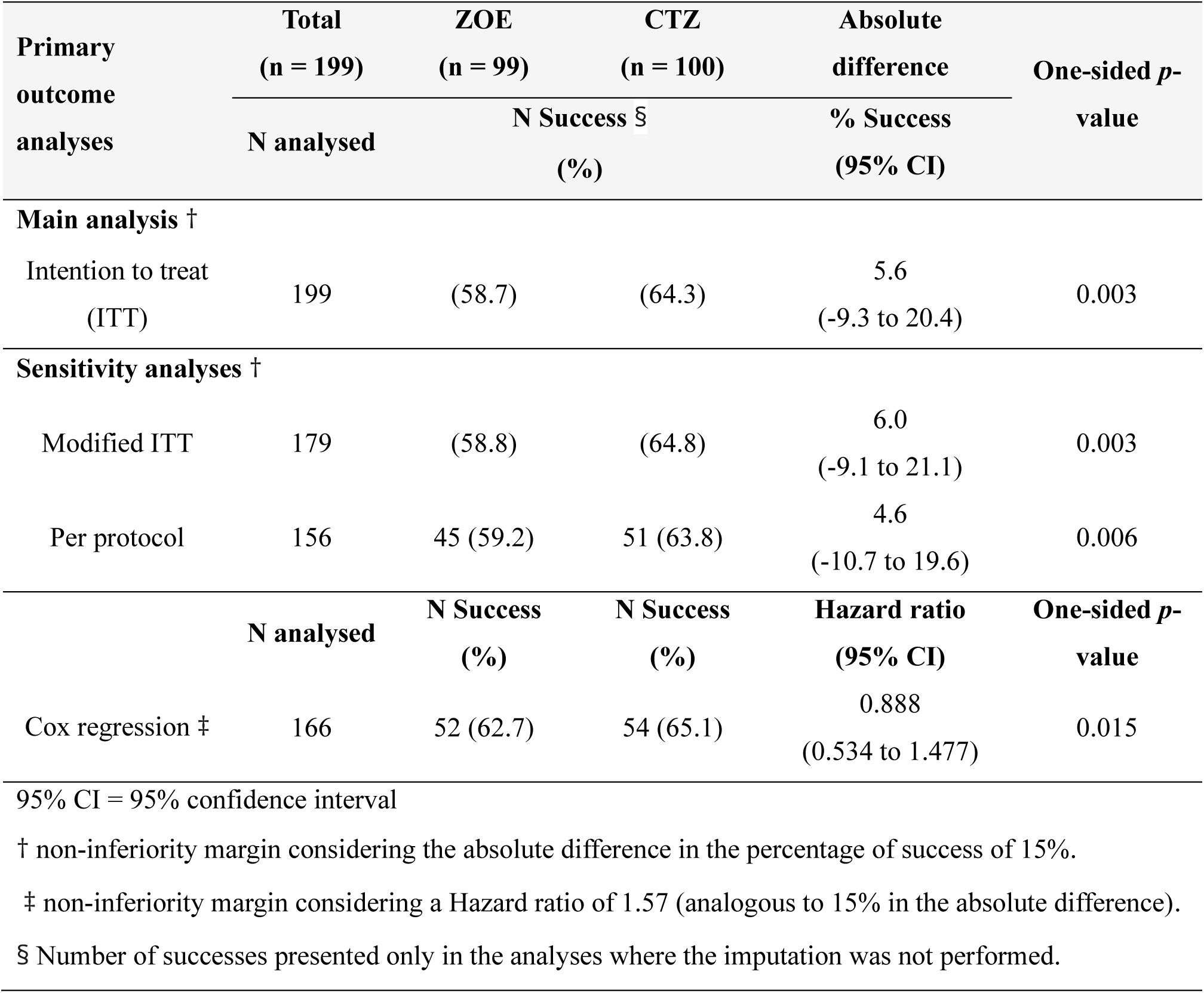
Main Intention-to-treat (ITT) analysis and sensitivity analyses using modified ITT (complete cases), per protocol, and Cox regression approaches comparing the overall success after 24 months of endodontically treated primary molars using the conventional pulpectomy technique (ZOE) and LSTR with CTZ paste (CTZ).

| **Primary outcome analyses** | **Total<br>(n = 199)** | **ZOE<br>(n = 99)** | **CTZ<br>(n = 100)** | **Absolute difference** | **One-sided *p*-value** |
|---|---|---|---|---|---|
|  | **N analysed** | **N Success** §<br>**(%)** |  | **% Success<br>(95% CI)** |  |
| **Main analysis** † |  |  |  |  |  |
| Intention to treat (ITT) | 199 | (58.7) | (64.3) | 5.6<br>(-9.3 to 20.4) | 0.003 |
| **Sensitivity analyses** † |  |  |  |  |  |
| Modified ITT | 179 | (58.8) | (64.8) | 6.0<br>(-9.1 to 21.1) | 0.003 |
| Per protocol | 156 | 45 (59.2) | 51 (63.8) | 4.6<br>(-10.7 to 19.6) | 0.006 |
|  | **N analysed** | **N Success<br>(%)** | **N Success<br>(%)** | **Hazard ratio<br>(95% CI)** | **One-sided *p*-value** |
| Cox regression ‡ | 166 | 52 (62.7) | 54 (65.1) | 0.888<br>(0.534 to 1.477) | 0.015 |

95% CI = 95% confidence interval
† non-inferiority margin considering the absolute difference in the percentage of success of 15%.
‡ non-inferiority margin considering a Hazard ratio of 1.57 (analogous to 15% in the absolute difference).
§ Number of successes presented only in the analyses where the imputation was not performed.

All sensitivity analyses provided consistent findings, supporting the robustness of the non-inferiority performance of the experimental treatment. The lower limit of the 95% CI was-9.1% and-10.7% in the modified ITT and per-protocol analyses, respectively (Table 2). The Cox regression analysis also supported non-inferiority. When applying a non-inferiority margin corresponding to an absolute difference of 15%, the one-sided *p*-value for the non-inferiority hypothesis remained below the predefined significance level of 5% (Table 2). Subgroup analyses likewise supported non-inferiority among females, children aged 3 to 5 years, treatments performed in second primary molars, and teeth presenting with abscess or fistula at baseline (Figure 2).

From the 156 children who completed the study, per-protocol clinical success was observed in 57/76 (75.0%) and 65/80 teeth (81.3%) in the ZOE and CTZ, respectively. Regarding per-protocol radiographic success, 54/76 teeth (71.1%) in the ZOE and 59/80 teeth (73.8%) in the CTZ showed no radiographic signs of treatment failure. In the full ITT analysis, both secondary outcomes showed a pattern consistent with that observed for the primary outcome of overall treatment success (Table 3).

**Table 3.** Secondary outcomes analyses of dichotomous variables comparing endodontically treated primary molars using the conventional pulpectomy technique (ZOE) and LSTR with CTZ paste (CTZ).

| **Secondary outcome analyses (dichotomous variables)** | **ZOE<br>(n = 99) †** | **CTZ<br>(n = 100) †** | **Absolute difference** | ***p*-value** |
|---|---|---|---|---|
|  | **% Success ‡** |  | **% Success<br>(95% CI)** |  |
| **Secondary outcomes related to the success (n recorded = 156)** |  |  |  |  |
| Clinical success | 74.2 | 81.0 | 6.8<br>(-6.6 to 20.3) | <0.001 * |
| Radiographic success | 71.2 | 75.1 | 3.9<br>(-9.5 to 17.2) | 0.003 * |
| **Secondary outcomes related to the postoperative assessment (n recorded = 177)** |  |  |  |  |
| Postoperative pain | 24.0 | 18.9 | -5.1<br>(-17.4 to 7.2) | 0.415 ** |
| Analgesic intake | 19.2 | 21.5 | 2.3<br>(-9.8 to 14.5) | 0.708 ** |
| Occurrence of swelling | 6.9 | 6.1 | -0.8<br>(-8.4 to 6.8) | 0.838 ** |

† n of randomized and analysed children (intention-to-treat – ITT - analyses).
‡ Percentages based on the ITT analysis using multiple imputation for missing outcome data.
\* one-sided p-value for non-inferiority hypothesis according to the Miettinen–Nurminen method stratified by the centre.
\*\* two-sided p-value derived by Wald approach stratified by the centre.

Postoperative pain and occurrence of swelling within the first 48 hours after treatment were evaluated as secondary outcomes. Parents were additionally asked whether their child required analgesic medication during the first 48 hours after treatment. No significant differences between groups were observed for any of these outcomes (Table 3).

Most children experienced little discomfort (score 1) immediately after treatment in both groups (62.1%; *n* = 110/177, 56 in ZOE and 54 in CTZ), with no statistically significant difference in mean scores between them. Likewise, child behaviour was predominantly definitely positive in both groups (57.6%; *n* = 102/177, 48 in ZOE and 54 in CTZ), with no statistically significant between-group differences. In contrast, the mean treatment time (SD) in the ZOE was 63.0 (16.3) minutes, approximately twice that in the CTZ, which averaged 36.3 (9.8) minutes; this difference was statistically significant (Table 4).

**Table 4.** Secondary outcomes analyses of ordinal and quantitative variables comparing endodontically treated primary molars using the conventional pulpectomy technique (ZOE) and LSTR with CTZ paste (CTZ).

| **Secondary outcome analyses (ordinal and quantitative variables)** | **ZOE**<br>**(n = 99) †** | **CTZ**<br>**(n = 100) †** | **Two-sided *p*-value \*** |
|  | **Mean (Standard Deviation)**<br>***Median (Interquartile Range)*** |  |  |
|---|---|---|---|
| Children’s behaviour | 3.3 (1.0)<br>4.0 (3.0 – 4.0) | 3.3 (1.0)<br>4.0 (3.0 – 4.0) | 1.000 |
| Children’s immediate discomfort | 1.7 (1.2)<br>1.0 (1.0 – 2.0) | 1.9 (1.4)<br>1.0 (1.0 – 2.0) | 0.464 |
| Clinical Time (min) | 63.0 (16.3)<br>61.3 (49.7 – 74.7) | 36.3 (9.8)<br>34.6 (28.4 – 42.9) | < 0.001 |

† n of randomized and analysed children (ITT analyses). The number of children recorded for these variables were 177.
\* two-sided p-value derived by Mann-Whitney test, pooled across imputations (n = 50) by Fisher’s method.

The reasons for treatment failure in each group are presented in Table 5. The development of a new radiolucent lesion or the progression of a pre-existing lesion was the most common cause of failure. Pathological external root resorption and the presence of abscess or fistula were also frequent findings associated with treatment failure. Apart from the postoperative events reported in Table 3, no additional adverse events were observed in either group.

**Table 5.** Reasons for treatment failures in endodontically treated primary molars using the conventional pulpectomy technique (ZOE), and LSTR with CTZ paste (CTZ).

| **Reasons for failures** | **ZOE** | **CTZ** |
|---|---|---|
|  | **N (%) †** |  |
| **Clinical Failures** |  |  |
| Abscess / Fistula | 12 (20.0) | 7 (13.7) |
| Pain | 5 (8.3) | 5 (9.8) |
| Restoration loss | 6 (10.0) | 7 (13.7) |
| **Radiographic failures** |  |  |
| Interradicular lesion<br>(new lesion or progression) | 17 (28.3) | 19 (37.3) |
| Internal resorption | 4 (6.7) | 4 (7.8) |
| External pathological resorption | 16 (26.7) | 9 (17.6) |
| **Total** | **60 (100.0)** | **51 (100.0)** |

† Number exceeds the total failures (31 for ZOE and 29 for CTZ) because some teeth presented more than one type of failure.

## 4 Discussion

Although endodontic treatment of primary teeth with pulp involvement improves children’s oral health-related quality of life compared with extraction,^6,7^ conventional pulpectomy is complex and requires considerable chair time.^24^ Consequently, simplified approaches, such as LSTR, may represent a viable alternative, provided that their success rates are not clinically inferior to those of conventional pulpectomy. We therefore conducted this multicentre randomised clinical trial evaluating the clinical and radiographic success of a LSTR using CTZ paste with that of conventional pulpectomy involving root canal instrumentation and ZOE filling in primary molars. The main finding was that LSTR with CTZ paste was non-inferior to conventional pulpectomy after 24 months of follow-up. Moreover, our secondary outcome analysis showed that LSTR required approximately half the chair time of conventional pulpectomy. To the best of our knowledge, this is the largest trial to date comparing LSTR with conventional endodontic treatment in primary teeth and the first to formally test a non-inferiority hypothesis.

Most previous studies comparing LSTR with conventional endodontic treatment had small sample sizes and relatively short follow-up periods, typically limited to 12 months.^25–28^ Consequently, the certainty of evidence supporting LSTR was rated as low in a recent systematic review.^2^ Furthermore, studies specifically evaluating CTZ paste have also been limited by small sample sizes and methodological shortcomings.^15–17^ Only one well-conducted randomised controlled trial has evaluated the long-term clinical and radiographic success of LSTR using CTZ paste compared with conventional pulpectomy using ZOE filling.^18^ The clinical protocols used in that trial and ours were closely comparable, with the same interventions and comparator. The main differences concerned the irrigating solution and restorative approach. Although our overall success rate was higher than that reported in the previous trial, the authors also observed similar success rates between groups at the 24-month follow-up. Moreover, the success rate observed in our study was comparable to that reported in the systematic review for LSTR using other antibiotic formulations. The review reported a pooled success rate of 53.9% at 24 months, based on three studies involving 154 teeth.^2^ Taken together, these findings suggest that our results are broadly consistent with previous evidence, while providing additional evidence from a trial specifically designed to assess non-inferiority.

Conventional pulpectomy is associated with high success and survival rates;^2,3^ however, its technical complexity, time-consuming nature, and dependence on patient cooperation may limit its implementation in daily paediatric clinical practice.^4,24^ In this context, LSTR offers a simplified alternative by eliminating the need for root canal instrumentation and allowing treatment to be concluded in a single visit.^2–4^ These characteristics may be particularly advantageous when treating young children, with limited cooperation or special healthcare needs, and in public oral health services, where access to the materials and equipment required for conventional pulpectomy may be limited.^5,24^ LSTR may therefore be a less costly and more accessible approach, as a cost-analysis study reported that LSTR with CTZ paste was 58.3% less expensive than conventional pulpectomy, considering dental materials, equipment depreciation, and professional chairside time.^5^ Its implementation in public health services could potentially expand access to endodontic treatment and help prevent the premature loss of primary teeth among children facing prolonged waiting times for specialised care.

LSTR with CTZ paste required approximately half the clinical time of conventional pulpectomy, corroborating previous findings of shorter treatment times with this approach.^24^ This difference is expected, given the additional time required for chemomechanical root canal preparation, including instrumentation, irrigation, and obturation. Despite this substantial reduction in treatment time, no significant differences were observed between groups in child behaviour during treatment, immediate postoperative discomfort, postoperative pain, or analgesic consumption, suggesting comparable patient-centred outcomes. Evidence in this area remains scarce, particularly for LSTR protocols using CTZ paste, and, to our knowledge, no previous studies have directly compared children’s behaviour during LSTR and conventional pulpectomy. Nevertheless, previous investigations evaluating different LSTR protocols also reported no significant differences in postoperative pain compared with conventional pulpectomy at 24 hours^29^ and two weeks,^30^ supporting the present findings. Collectively, these results suggest that the simplification of the endodontic procedure achieved through LSTR does not appear to compromise patient-centred outcomes.

The presence of tetracycline in the CTZ paste represents a limitation of the presented LSTR protocol. Tooth discolouration is a well-documented adverse effect; therefore, LSTR protocols incorporating tetracyclines are more appropriate for primary molars and should be avoided in anterior teeth.^18,24^ In addition, antibiotic-containing formulations may be associated with sensitisation and hypersensitivity reactions, and antibiotic resistance is a concern.^11^ However, a systematic review evaluating antibiotic mixtures used in LSTR reported that none of the included studies identified sensitivity-related adverse events, regardless of whether tetracyclines were included.^11^ Nevertheless, further research investigating potential substitutes for tetracycline in CTZ paste is warranted.

The present study has methodological limitations, the main one being the discontinuation of one participating centre. However, the ITT analysis had been prespecified as the primary approach in the study protocol, and sensitivity analyses were also performed, excluding participants from the discontinued centre and restricting the analysis to the per-protocol population. Non-inferiority was demonstrated consistently across all analyses, suggesting that the discontinuation of this centre did not materially alter the main study findings.

Despite its limitations, this study has notable strengths. The multicentre design enhances the external validity and generalisability of the findings by assessing interventions across different clinical settings and populations. Furthermore, the 24-month follow-up period provides valuable evidence in an area where such data remain limited. The non-inferiority framework is also a key strength, as it is particularly appropriate for evaluating a simplified treatment approach that may offer practical advantages over conventional therapy. Finally, the consistency of findings across the prespecified analyses further supports the non-inferiority conclusion.

In conclusion, the LSTR technique with CTZ paste is non-inferior to the conventional pulpectomy technique using ZOE as filling material in the treatment of necrotic primary molars after 24 months of follow-up. This finding should be interpreted in the context of the standardised conditions of our trial, including strict diagnostic and eligibility criteria and a standardised treatment protocol. Further pragmatic trials are therefore needed to determine whether these findings can be replicated under routine clinical conditions.

## Statements

### Data availability statement

The pseudo-anonymised dataset (https://repositorio.uspdigital.usp.br/handle/item/867), analysis codes (https://repositorio.uspdigital.usp.br/handle/item/888) and full statistical report (https://repositorio.uspdigital.usp.br/handle/item/887) are available in the public data repository of the University of São Paulo.

### Ethics approval statement

The study protocol was reviewed and approved by the local ethical committees from the enrolled centres: the Schools of Dentistry of the *Universidade de São Paulo* (USP), *Universidade Federal de Santa Catarina* (UFSC), *Universidade do Estado do Rio de Janeiro* (UERJ) and *Universidade Iguaçu* (UNIG), under the approval number 42602820.0.1001.0121.

### Patient consent statement

All parents or legal guardians of the participants signed an informed consent form, and the participants signed an assent form.

## Data Availability

The pseudo-anonymised dataset, analysis codes and full statistical report are available in the public data repository of the University of Sao Paulo.

https://repositorio.uspdigital.usp.br/handle/item/867

https://repositorio.uspdigital.usp.br/handle/item/888

https://repositorio.uspdigital.usp.br/handle/item/887

